# Solving the “right” problems for effective machine learning driven in vitro fertilization

**DOI:** 10.1101/2021.10.07.21264503

**Authors:** Itay Erlich, Assaf Ben-Meir, Iris Har-Vardi, James A. Grifo, Assaf Zaritsky

## Abstract

Automated live embryo imaging has transformed in-vitro fertilization (IVF) into a data-intensive field. Unlike clinicians who rank embryos from the same IVF cycle cohort based on the embryos visual quality and determine how many embryos to transfer based on clinical factors, machine learning solutions usually combine these steps by optimizing for implantation prediction and using the same model for ranking the embryos within a cohort. Here we establish that this strategy can lead to sub-optimal selection of embryos. We reveal that despite enhancing implantation prediction, inclusion of clinical properties hampers ranking. Moreover, we find that ambiguous labels of failed implantations, due to either low quality embryos or poor clinical factors, confound both the optimal ranking and even implantation prediction. To overcome these limitations, we propose conceptual and practical steps to enhance machine-learning driven IVF solutions. These consist of separating the optimizing of implantation from ranking by focusing on visual properties for ranking, and reducing label ambiguity.

**Lay Summary:** *Background:* In vitro fertilization (IVF) is the process where a cohort of embryos are developed in a laboratory followed by selecting a few to transfer in the patient’s uterus. After approximately forty years of low-throughput, automated live embryo imaging has transformed IVF into a data-intensive field leading to the development of unbiased and automated methods that rely on machine learning for embryo assessment. These advances are now revolutionizing the field with recent retrospective papers demonstrating computational models comparable and even exceeding clinicians’ performance, startups and medical companies are securing significant funds and at advanced stages of regulatory approvals. Traditionally, embryo selection is performed by clinicians ranking cohort embryos based solely on their visual qualities to estimate implantation potential, and then using non-visual clinical properties that are common to all cohort embryos to decide how many embryos to transfer. Machine learning solutions usually combine these two steps by optimizing for implantation prediction and using the same model for ranking the embryos within a cohort under the implicit assumption that training to predict implantation potential also optimizes a solution to the problem of ranking embryos from a specific cohort.

*Results:* In this multi-center retrospective study we analyzed over 48,000 live imaged embryos to provide evidence that the common machine-learning scheme of training a model to predict implantation and using the same model for embryo ranking is wrong. We made this point by explicitly decoupling the problems of embryo implantation prediction and ranking with a set of computational analyses. We demonstrated that: (1) Using clinical cohort-related information (oocyte age) improves embryo implantation prediction but deteriorates ranking, and that (2) The label ambiguity of the embryos that failed to implant (it is not known whether the embryo or the external factors were the reason for failure) deteriorates embryo ranking and even the ability to accurately predict implantation. Our study provides a quantitative mapping of the tradeoffs between data volume, label ambiguity and embryo quality. In a key result, we reveal that considering embryos that were excluded based on their poor visual appearance (called discarded embryos), although commonly thought as trivially discriminated from high quality embryos, enhances embryo ranking by reducing the ambiguity in their (negative) labels. These results establish the benefit of harnessing the availability of extensive data and reliable labels in discarded embryos to improve embryo ranking and implantation prediction.

*Outlook:* We make two practical recommendations for devising machine learning solutions to embryo selection that will open the door for future advancements by data scientists and IVF technology developers. Namely, training models for embryo ranking should: (1) focus exclusively on embryo intrinsic features. (2) include less ambiguous negative labels, such as discarded embryos. In the era of machine learning, these guidelines will shift back the traditional two-step process of optimizing embryo ranking and implantation prediction independently under the appropriate assumptions - an approach better reflecting the clinician’s decision that involves the evaluation of all the embryos in the context of its cohort.

## Introduction

One of the major challenges in achieving effective in vitro fertilization (IVF) is selecting the specific embryos for implantation from a cohort of “sibling” embryos from the same IVF cycle [1, 2]. The decision as to how many and which embryos to implant back to the uterus is driven by the goal of achieving the birth of exactly one healthy baby [3, 4]. Toward this goal, clinicians score the visual properties of the developing embryos, alongside non-visual clinical parameters that are common to all the embryos from the same cohort, such as previous IVF outcomes, oocyte age and underweight/obesity to estimate the probability of a successful implantation and use these parameters to decide how many and which embryos to implant [2, 5, 6, 7, 8] (Fig. 1A). Traditionally, embryo grading and selection are subjectively performed in a two-step process where clinicians first rank cohort embryos based solely on their visual qualities, as a proxy for embryo implantation potential, and then use non-visual clinical properties as a proxy for the embryo-extrinsic factors involved in implantation, to decide how many embryos to transfer [9, 7]. Recent advances in automated imaging and computation have led to objective and systematic data-driven approaches for embryo evaluation, where machine learning models are trained to output a score that predicts implantation potential [10, 11, 12, 13, 14, 15, 16]. These scores are then used to rank the embryos in an IVF cycle cohort and assist the clinician in making decisions regarding which embryos should be transferred. This approach of first using massive datasets to train a classifier that predicts retrospective implantation outcomes, and then using its prediction to rank embryos for transfer is performed under the implicit premise that a model trained to predict implantation potential is suitable for ranking embryos from a specific cycle cohort.

**Figure 1:**
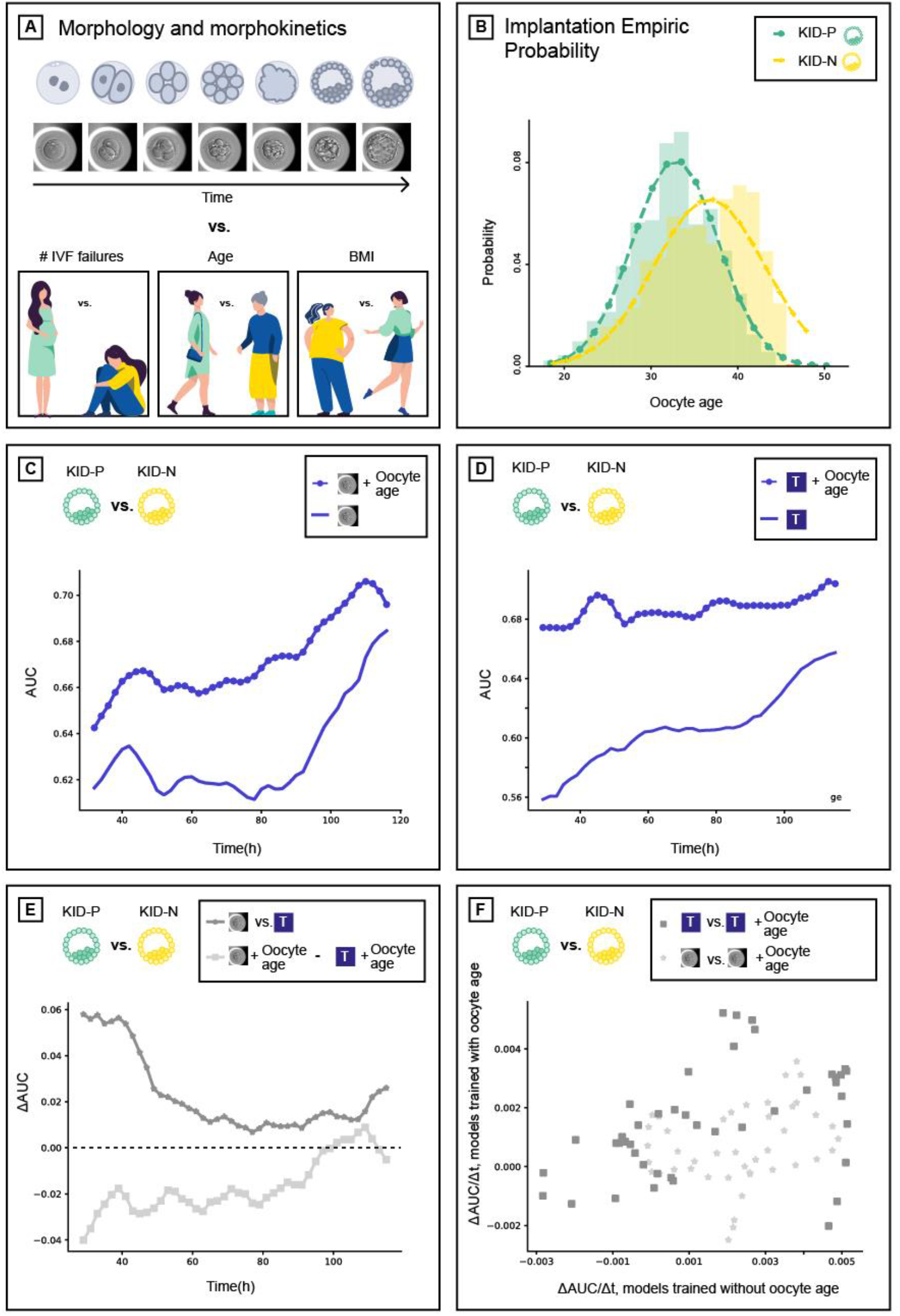
Decoupling embryo scoring and ranking using intrinsic and extrinsic features. (**A**) Embryo implantation potential relies on the embryo’s visual and clinical properties. (**B**) Distribution of implanted (KID-P) versus non-implanted (KID-N) embryos according to oocyte age. The average oocyte age of the KID-P (KID-N) embryos was 32.82 (36.74) with a standard deviation of 4.93 (6.33). N KID-P = 1694, N KID-N = 7122. (**C-D**) Comparison of the implantation prediction performance (AUC) of models trained with or without the oocyte age over time since fertilization. (**C**) Embryo morphological features (marked with an embryo image). (**D**) Embryo morphokinetic features (marked with “T”). (**E**) Comparing implantation prediction performance: the morphology-based classifier outperformed morphokinetic-based classifier but morphokinetic and oocyte age outperformed morphology and oocyte age. (**F**) Correlation between the temporal derivative of the morphology-based classification with and without oocyte age (Pearson correlation R = 0.43) versus correlation between the temporal derivative of the morphokinetic-based classification with and without oocyte age (Pearson correlation R = 0.27).

Here, we challenge this axiom. There is a general consensus that non-visual clinical properties contribute to the prediction of implantation outcomes [17, 18, 6, 14], but that such information is not relevant for ranking embryos in the context of a single IVF cycle cohort that share the same non-visual clinical properties. This implies that the optimization task solved through machine learning in practice is different from the actual decision made in the clinic. Moreover, while successful implantation inherently implies that an embryo is of high implantation quality, a failed implantation can be caused by a defective embryo or be related to poor maternal clinical factors (such as oocyte age, uterus cavity and receptivity of the endometrium). This creates uncertainty in the ground truth labels of failed implanted embryos that may hamper the ability of machine learning models to generalize well during training. It is not clear whether using machine learning models trained with clinical information can hamper the selection of the most promising embryos for implantation, and what is the magnitude of the errors caused by the ambiguous labels of failed implantations.

We hypothesized that predicting implantation is not the optimal machine learning strategy for solving the embryo-ranking problem because features derived from clinical information shared among embryos from the same cohort are not relevant for the ranking task and because ambiguous labels confound the optimal ranking. We confirmed this hypothesis by comprehensively assessing the contribution of visual embryo properties and clinical factors, and by evaluating several criteria for training data selection with less ambiguous labels. Finally, we quantified the extent of these non-optimal strategies and propose conceptual and practical setups to overcome these limitations by training and applying separate classifiers for the different tasks of predicting embryo implantation potential and ranking.

## Results

### Decoupling embryo scoring and ranking using embryo morphology, morphokinetics and oocyte age

One important application of machine learning in IVF clinics is the prediction of successful implantation. This task involves training a machine learning classifier using a large number of embryos that either succeeded (KID-positive, KID-P) or failed (KID-negative, KID-N) to implant back to the uterus (Methods). During classifier training, each embryo is represented by its visual properties and non-visual clinical properties that are provided together with the implantation outcome as the target label (Fig. 1A). The trained classifier receives as input new embryos and predicts a score corresponding to their implantation potential; i.e., the probability for successful implantation. The visual embryo properties include the appearance of the embryo (termed morphology) and the time it takes to reach predefined stages in development (termed morphokinetics). Oocyte age, which in this study is equivalent to the maternal age (see Methods) is the most studied non-visual clinical property correlating with implantation. Other non-visual clinical factors may include infertility causes, history of previous IVF treatments, uterus cavity, receptivity of the endometrium, or any other property that is shared across all the embryos from the same cohort and thus cannot be directly extracted from the specific embryo’s image. The embryo-specific visual properties reflect the intrinsic embryo’s potential for implantation, whereas the cohort-specific non-visual clinical properties correspond to the embryo-extrinsic parameters. Successful implantation is heavily dependent on both the embryo-specific visual appearance and on the cohort-specific non-visual clinical factors.

We sought to systematically assess the contribution of non-visual clinical properties to embryo implantation by comparing machine learning models that rely on visual embryo features with or without non-visual clinical properties. Our data were composed of time-lapse videos collected from 4 clinics: 7904 IVF cycles that included 1314 KID-P and 6485 KID-N for Train, and 380 KID-P and 637 KID-N for Test (Methods, Table S1). The non-visual clinical property chosen was the oocyte age, shared among all embryos from the same IVF cycle cohort, and is probably the most common embryo-extrinsic property used for the embryo implantation prediction task [17]. First, we validated that the KID-P embryos were associated with younger oocytes (Fig. 1B). Next, we preprocessed the images (Fig. S1) using the morphology-based deep learning classifier described in a companion paper and in the Methods (Fig. S2A). This classifier provides a continuous score reflecting the implantation score for each embryo over time with or without the oocyte age as additional input to the embryo image (Methods, Fig. S2C). As expected, the classifier that had access to oocyte age outperformed the classifier that relied on morphology alone over time (Fig. 1C). To further verify that the oocyte age contributes to visual properties we devised a new classifier that calculated a continuous implantation score from the embryo morphokinetic properties (Fig. S2B, Methods). The timing of a set of fifteen hallmark morphokinetic events were identified automatically by combining machine-learning and dynamic programming (Figs. S3-4). These morphokinetic features were used to train a deep learning classifier to predict the implantation potential of an embryo based on its developmental history (see Methods for full details). Including the oocyte age as an additional feature improved the morphokinetic performance over time (Fig. 1D). Although the morphology-based classifier was more accurate than the morphokinetic-based classifier, the inclusion of oocyte age flipped the classification performance in favor of the morphokinetic model (Fig. 1E). This result suggests that the residual contribution of oocyte age to morphokinetic features was greater than the contribution of oocyte age to morphological features in the context of the implantation classification task. Correlating the temporal derivative of the corresponding classifier pairs showed that the temporal trend of the classification performance was more similar between the classifier pair that was trained with morphological features (with and without the oocyte age) than the classifier trained with the morphokinetic features, thus providing further evidence that the common classification-related information between morphology features and oocyte age was higher than between morphokinetic features and oocyte age (Fig. 1F). Since oocyte age is common to all cohort embryos and thus irrelevant for the ranking task, the prediction of the higher performing classifier for the implantation potential is not necessarily aligned with the clinician’s task of ranking “sibling” embryos originating from the same IVF cycle. Hence, although the classifier trained with both morphokinetic features and oocyte age outperformed the classifier trained with both morphological features and oocyte age (especially in early embryo development), embryo morphological features seem more appropriate for the task of ranking high quality embryos. Thus, beyond providing a quantitative assessment of the contribution of oocyte age to implantation prediction, these results raise the possibility that clinical properties that are common across embryos from the same cohort contribute to the task of embryo scoring toward implantation but are not the optimal properties for the task of embryo ranking.

### Optimizing features for prediction of embryo implantation is not an optimal strategy for the task of embryo ranking

Embryos in an IVF cycle are partitioned into three groups by the clinician based on their visual properties. These are composed of 1) transferred embryos - the most promising embryos for implantation (Video S1), 2) frozen embryos - other visually high-quality embryos that are kept for transfers in future IVF cycles and 3) discarded embryos - those selected to be excluded based on their poor visual appearance (Video S2-S4). Discarded embryos can be further partitioned into two subgroups: those that reached blastulation (termed *blastocyst-discarded*, Video S2) and those that did not reach blastulation (termed *underdeveloped-discarded*, Videos S3-4). These partitions define the expected ordered ranking of embryos based on their visual appearance (Fig. 2A).

**Figure 2:**
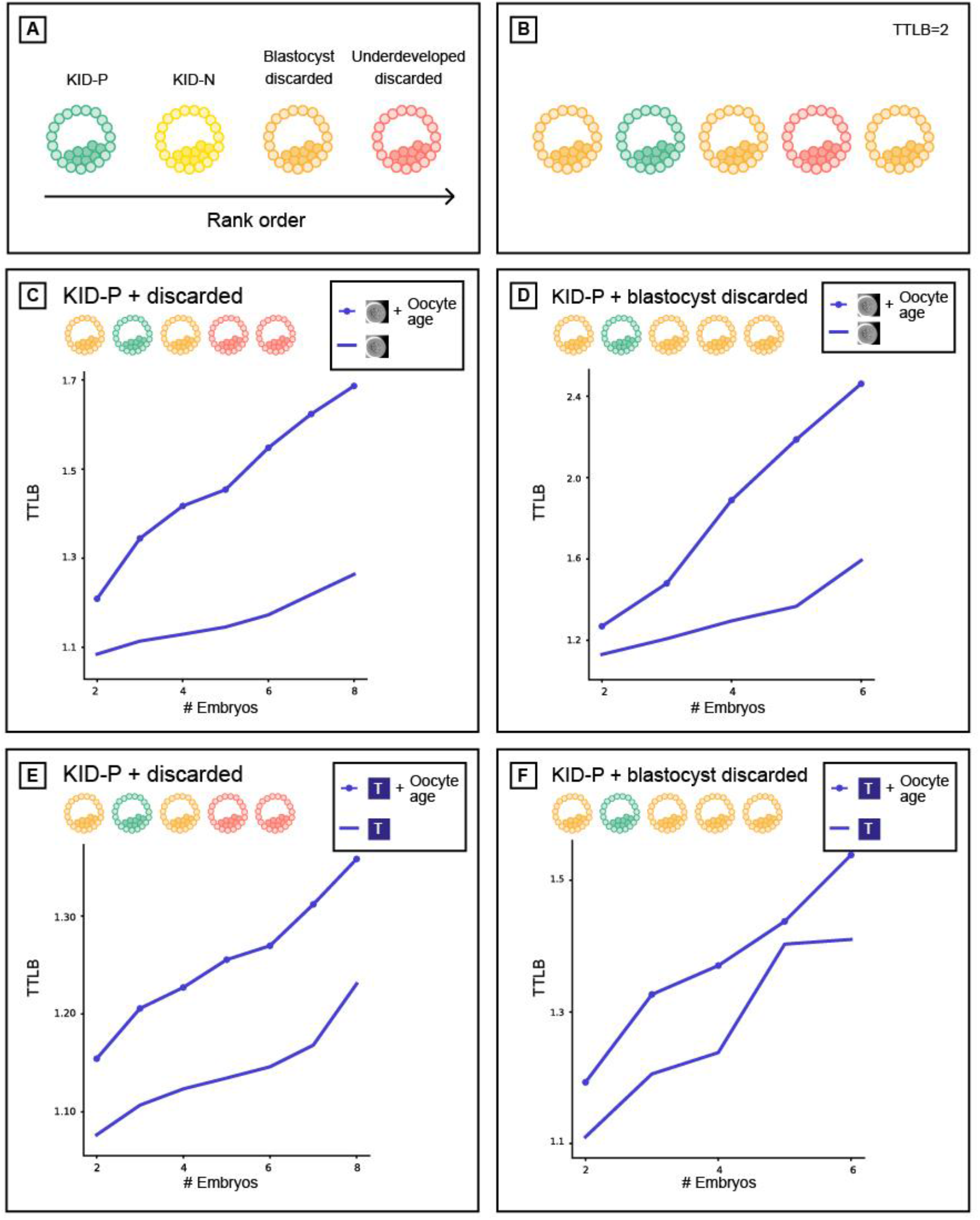
Using oocyte age as feature for improving implantation prediction reduced the accuracy of ranking embryos from the same cohort. (**A**) Embryo visual evaluation: KID-P > KID-N > Blastocyst-discarded > underdeveloped-discarded. KID-N and frozen embryos are not considered in embryo ranking evaluations because there is no clear distinction based on their visual properties. (**B**) Time To Live Birth (TTLB) for sub-cohorts of sibling embryos, as a function of the number of embryos considered for each cohort. Each sibling sub-cohort consisted of one implanted embryo and multiple discarded embryos sampled from the full cohort. In the example here the number of embryos was 5 and the TTLB was 2 because the KID-P embryo was ranked second. (**C-D**) Morphology-based models (marked with an embryo image) with or without oocyte age. (**C**) Ranking KID-P and all discarded embryos. Number of cycles with n = 2-8 embryos: 201, 180, 163, 141, 115, 93, 67. Corresponding Wilcoxon signed rank tests on the null hypothesis that the two ranking schemes were drawn from the same matched distribution: <0.0001, <0.0001, <0.0001, <0.0001, <0.0001, <0.0001, 0.005. (**D**) Ranking KID-P and blastocyst-discarded embryos. Number of cycles with n = 2-6 embryos: 145, 98, 54, 32, 13. Corresponding Wilcoxon signed-rank tests: <0.0001, <0.0001, <0.0001, 0.001, 0.074. (**E-F**) Morphokinetic-based models (marked with “T”) with or without oocyte age. Number of cycles according to panels C-D respectively. (**E**) Ranking KID-P and all discarded embryos. Wilcoxon signed-rank tests: 0.001, 0.001, 0.001, 0.001, 0.004, 0.004, 0.039. (**F**) Ranking KID-P and blastocyst-discarded embryos. Wilcoxon signed-rank tests: 0.004, 0.004, 0.071, 0.157, 0.655.

A basic expectation of an embryo-ranking model is to prioritize KID-P over discarded embryos. To assess the performance of models trained to predict implantation results (KID-P versus KID-N) on the task of embryo ranking, we evaluated the average *time to live birth* (TTLB), the number of attempts for embryo transfer from an IVF cycle until a successful implantation was reached, where the transfer order was determined by the model’s ranking based on its classification score [19]. For example, a TTLB value of 2 implies a cohort where the KID-P was ranked second; i.e., one discarded embryo had a higher classification score (Fig. 2B). The analysis included embryo cohorts that contained a successful implantation (KID-P) and multiple discarded embryos. While it is widely believed that transferring a discarded embryo will result in a failed implantation, the implantation outcome for frozen embryos is not clear-cut; hence, they were not considered in this analysis. To avoid the bias of cohorts with different numbers of embryos we compared the average TTLB of sub-cohorts consisting of the same number of embryos sampled from the full cohorts (Methods).

Oocyte age contributed to improved implantation prediction (Fig. 1C-D) but this information is not relevant for ranking embryos from the same cohort that share the same oocyte age. To test the hypothesis that optimizing features for the KID-P versus KID-N classification task can deteriorate ranking accuracy, we compared the performance of two classifiers that were trained with morphological features with or without the oocyte age. We first ranked sub-cohorts of one KID-P and multiple discarded embryos (Methods). The average TTLB was at most 1.25 and the inclusion of oocyte age led to less efficient (i.e., higher TTLB) ranking, that required on average 1.7 attempts to achieve successful implantation (Fig. 2C). The same less efficient ranking with oocyte age was observed for sub-cohorts with one KID-P and multiple blastocyst-discarded embryos (Fig. 2D). As expected, ranking with blastocyst-discarded embryos constituted a more challenging task because the visual defects are less obvious with respect to embryos that did not reach blastulation. Similar trends were obtained for morphokinetic features with or without oocyte age (Fig. 2E-F). Altogether, these results suggest that selecting features to optimize implantation prediction is not the optimal strategy for embryo ranking, specifically when these features are clinical properties that are common to all embryos in a cohort.

### KID-N embryos are not the ideal negative labels for the task of embryo ranking

After demonstrating that the features selected for their performance at implantation prediction were not optimal for embryo ranking we asked how different training data would affect the models’ performance when discriminating high quality from poor quality embryos. We defined a set of classification tasks that consisted of discriminating KID-P embryos from different subsets of discarded and/or unsuccessful implanted embryos. First, discriminating KID-P from all discarded embryos (Fig. 3A). This is an easy classification task because discarded embryos are characterized by evident visual defects. The second classification task involved discriminating KID-P from blastocyst-discarded embryos (Fig. 3B). The third involved discriminating KID-P from a subset that included both the KID-N as well as the discarded embryos (Fig. 3C). This latter setting is the one most similar to the true embryo ranking tasks in the clinic. For each of these embryo subsets we trained six morphology-based classifiers (Table S2) and their classification performance was compared on different classification tasks (Table S3, Fig. 3A-C).

**Figure 3:**
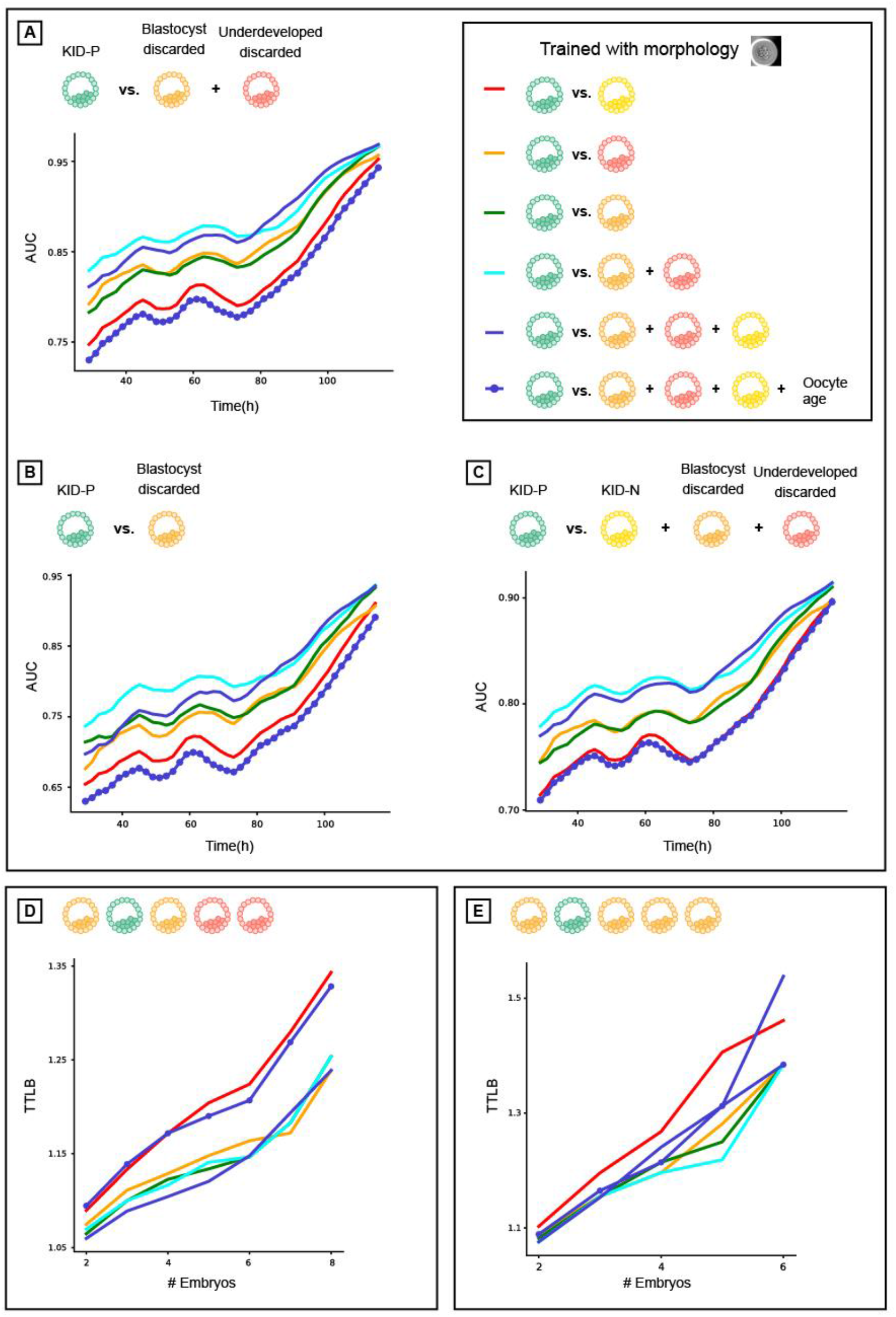
KID-N embryos are not the ideal negative label for solving the task of embryo ranking. (**A-C**) Prediction accuracy (AUC) over time since fertilization (higher scores reflect better performance). (**D-E**) Time To Live Birth (TTLB) for cohorts of sibling embryos, as a function of the number of embryos in each cohort (lower scores reflect better performance). Each siblings sub-cohort consisted of one implanted embryo and multiple discarded embryos from the same cohort. Number of cycles corresponding to those reported in Fig. 2. (**A-E**) All models were trained with morphological features. (**A**) KID-P versus discarded. (**B**) KID-P versus blastocyst-discarded. (**C**) KID-P versus KID-N and discarded. (**D**) Ranking KID-P and all discarded embryos. (**E**) Ranking KID-P and blastocyst-discarded embryos.

Classifiers trained to predict implantation (KID-P versus KID-N) were less accurate than the classifiers that were trained with discarded embryos on all tasks (Fig. 3A-C). Inclusion of the oocyte age deteriorated the performance to the levels of the classifiers that were trained to predict implantation (Fig. 3A-C). Direct evaluation of time to live birth established that models trained to predict embryo implantation were not necessarily optimal for the task of embryo ranking (Fig. 3D-E).

The KID-P versus KID-N labeled training data was severely limited in size because only a small subset of all embryos were selected for transfer and thus discarded embryos constitute the vast majority of embryos in IVF clinic in our study (Table S1) and in general [13, 20]. To assess the contribution of the extended volume of training data we limited the amount of negative labeled training data across the different categories of discarded embryos to align with the number of KID-N embryos (Table S2) and demonstrated that classifiers trained with discarded embryos outperformed the classifier trained to predict implantation even without extra training data (Fig. 3A-E, compare orange and green versus red). Additional training data further enhanced the classifiers’ performance (Fig. S5). Similar results were obtained when using morphokinetic features (Figs. S6-7). Altogether, these results suggest that the elementary requirement of distinguishing poor from high quality embryos broke more frequently in the KID-P/KID-N trained classifiers, suggesting that KID-P/KID-N are not the optimal training data for the task of embryo ranking.

### Tradeoffs between training labels ambiguity and specificity in predicting implantation and ranking

In the clinic, embryo ranking within an IVF cycle is exclusively determined by assessing the embryos’ visual properties. In contrast, in the machine-learning solution, visual embryo properties only provide partial information in the context of implantation prediction. Failed implantations can be caused either by defected embryos or by embryo-extrinsic maternal clinical properties leading to non-receptive endometrium. This limitation translated to ambiguous (noisy) KID-N labels that can mislead the classifier during model training. The confidence in discarded embryos labels, in the context of successful or failed implantation is high because the criterion of being discarded is inherently visual and thus is not/less affected by external factors.

To assess the contribution of training data volume and confidence in labels we turned again to the task of predicting implantation. We found that including the discarded embryos as negative labels in addition to KID-N during training, enhanced the classifier’s performance in the KID-P versus KID-N classification task (Fig. 4A). Similar results were achieved when using morphokinetic features (Fig. S8). These results imply that despite the fact that the KID-P/KID-N classifier was trained for the most visually challenging task, the availability of extensive data and reliable labels improved classification performance.

**Figure 4:**
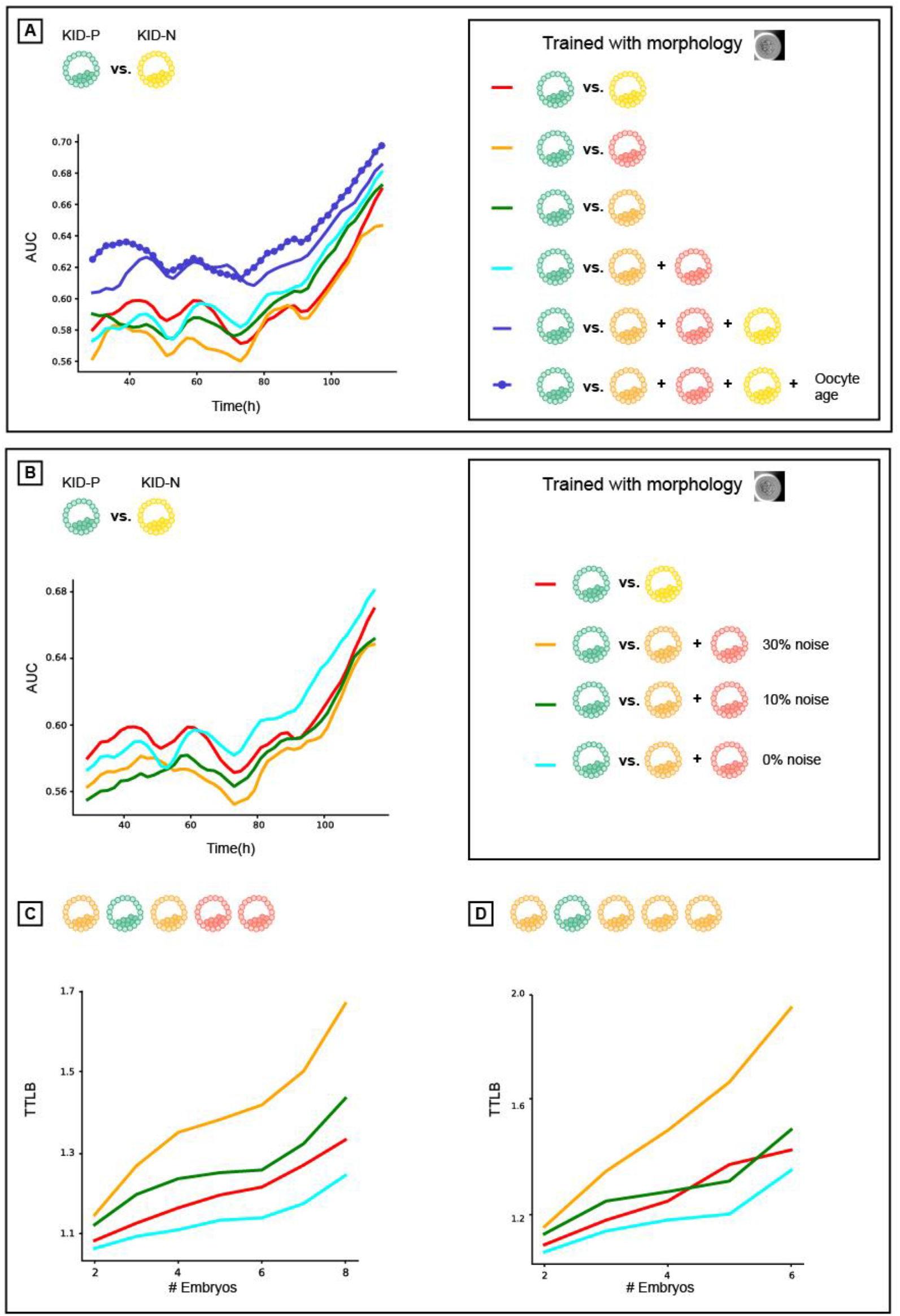
Analysis of implantation prediction and ranking performance as a function of negative label ambiguity and label specificity. All classifiers were trained using morphological features. (**A**) AUC of implantation prediction (KID-p versus KID-N) over time. Comparison of models trained to discriminate KID-P from different sets of negative labels: KID-N, discarded, blastocyst discarded, underdeveloped discarded, and KID-N+discarded with or without oocyte age. (**B**) AUC of implantation prediction (KID-p versus KID-N) over time for models trained to discriminate between KID-P and discarded embryos with increasing fractions of label ambiguity - flipping “KID-P” to “discarded” labels (see Methods). (**C-D**) Time To Live Birth (TTLB) for models trained in panel B. (**C**) KID-P versus all discarded. (**D**) KID-P versus blastocyst-discarded.

The contribution of training label specificity to a particular classification task was demonstrated by the superiority of the model trained with the KID-N and the discarded embryos over the model trained with discarded embryos alone (Fig. 4A – blue versus cyan correspondingly, both have comparable volume of negative training labels). This result is strengthen by the lack of a similar trend in other classification tasks where the addition of KID-N embryos to the training set did not contribute much to the performance (Fig. 3). These data demonstrate the tradeoff between label ambiguity and specificity and reveal that the combination of reduced ambiguity (but reduced specificity) in the discarded embryo labels along with increased specificity (but increased ambiguity) in the KID-N labels led to the most accurate implantation predictions.

To identify the breaking point between label ambiguity and specificity we simulated a gradual deterioration in the negative labels by flipping increasing fractions of positive labels to negative labels. This mimics the ambiguity of KID-N labels, where some viable embryos fail to implant for extrinsic maternal reasons and are thus mistakenly labeled as negatives. We evaluated the optimization problem of training models to discriminate KID-P from discarded embryos because of the higher confidence in the discarded embryos’ negative label. This analysis revealed that flipping 10% of the discarded embryo labels was sufficient to drop the performance of the KID-P/discarded trained classifier below that of the KID-P/KID-N-trained classifier for the tasks of predicting implantation potential and ranking, thus underscoring the fact that small fractions of erroneous labels can cause major deteriorations in the model’s performance (Fig. 4B-D). Altogether, these results highlight the interplay between label ambiguity, label specificity to a particular classification task and data volume during training in the context of IVF (Fig. 5).

**Figure 5:**
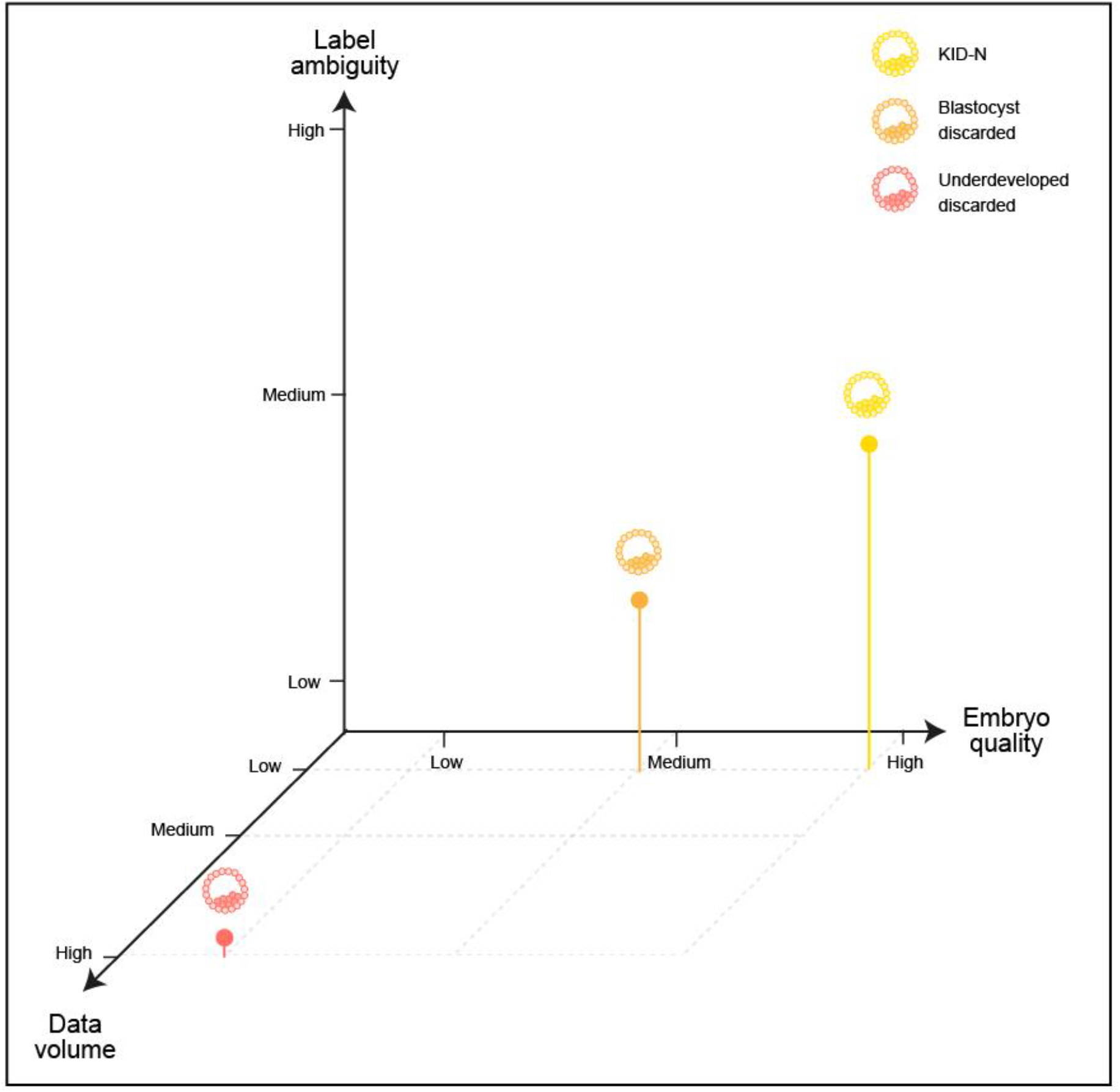
Trade-offs between data volume, label ambiguity and specificity to a particular classification task of the negative labels. In our case the specificity was aligned with the embryo quality (KID-P > blastocyst-discarded > underdeveloped-discarded, Fig. 2A). Data volume of underdeveloped-discarded was the highest (Table S1). Label ambiguity was high for KID-N. The positive labels were always KID-P.

## Discussion

There is a consensus that clinical properties that are shared across all embryos within a cohort, such as oocyte age, play a key role in determining embryo implantation potential but have no effect on embryo ranking within a cohort. It is also well-accepted that some KID-N embryos are functional embryos that failed implantation due to embryo-extrinsic clinical factors. In this study we took the next conceptual step by demonstrating that training classifiers to predict implantation can lead to non-optimal transfer decisions as evident by the higher probabilities of preferring obviously visually defective embryos over successfully implanted embryos.

These results raise concerns as to the current common pipeline for machine-learning driven IVF that directly imply several practical recommendations. First, training models for embryo ranking should focus exclusively on embryo intrinsic features. These may include engineered features that measure specific embryo properties such as the appearance of pronuclei and fading timing (tPNa, tPNf), the number of pronuclei, pronuclei shape, symmetry [21, 22, 23] blastocyst expanded diameter and trophectoderm cell cycle length [24], temporal events [25, 26] and/or training deep neural networks on the raw images [13] while avoiding embryo-extrinsic clinical factors that are shared by the other sibling embryos within the cohort. Second, the ambiguity in the KID-N labels makes implantation prediction a sub-optimal optimization problem both in model training as well as in model evaluation. Thus training with less ambiguous negative labels, such as discarded embryos, can enhance ranking accuracy (and perhaps even implantation prediction). Training with less ambiguous negative labels can either be implemented by including discarded embryos in the training binary classification tasks, as demonstrated here, and/or by defining optimization problems that are more specific for ranking [27]. The current data suggest that this approach can be beneficial using discarded embryos as negative labels even though discarded embryos are commonly thought as trivially discriminated from high quality embryos (low label specificity). Beyond better reflecting the clinician’s decision that involves the evaluation of all the embryos in an IVF cycle, another benefit of considering discarded embryos lies in the magnitude of data available for training that exceeded five-folds in the current dataset, and bypasses the biased selection of transferred embryos.

Hence avoiding/reducing the confounding effects of embryo-extrinsic clinical factors and ambiguous labels, when solving a ranking problem, as well as using all the available embryos to increase training data, are key practical concepts to consider when devising machine learning solutions for IVF. We hope that the awareness raised by our findings will bring back the traditional two-step process of ranking embryos within a cohort and determining how many embryos to transfer based on embryo scoring that approximates implantation probabilities, where each of these tasks is optimized independently under the appropriate assumptions.

## Methods

### Experiments

#### Data collection and ethics

The data were retrospectively collected from four clinics: the Ein-Kerem, and Mt. Scopus campuses of the Hadassah Hebrew University Medical Center, the Soroka University Medical Center, and the NYU Langone Prelude Fertility Center. After fertilization, the embryos were incubated in EmbryoscopeTM (Vitrolife, Copenhagen, Denmark), a time-lapse incubation and imaging system that acquires seven-layers of z-stack images 15µm apart at each time point every 15 to 20 minutes, where time 0 is defined as the fertilization time. We used the central focal plane, which was usually the most focused, for further analysis, on a total of roughly 360-480 images over a 5-day period for a single embryo. Time-lapse sequences of embryos were collected between July 2014 and December 2019 for oocytes aged 23.6-43.6 years (Fig. 1B). This study did not include donor oocyte IVF sessions and transfer of previously frozen embryos and thus the oocyte age is identical to the patient/maternal age and is shared among all embryos in a cohort. Embryos that were discarded before 114 hours were excluded from further analysis. Human embryo image/video data collected from patients were used in this study with institutional review board approval from the Investigation Review Board of Hadassah Hebrew University Medical Center (IRB# HMO-006-20). Overall, our dataset contained 47162 recorded videos of embryos collected from 7904 patients with informed consent for research and publication, under an institutional review board approval for secondary research use. Table S1 details the data splits between train and test.

#### Annotation of embryo clinical quality

Once the embryos reached the desired developmental stage, single or more embryos from a cohort were transferred. For those embryos that were transferred, implantation tagging was defined by one of the three possible Known Implantation Data (KID) tags: (i) KID-positive (KID-P): when all transferred embryos were successfully implanted (Video S1); (ii) KID-negative (KID-N): when none of the transferred embryos were successfully implanted; (iii) KID-unknown (KID-UK): when the outcome of each transferred embryo was uncertain. KID-UK embryos were excluded from further analysis due to their ambiguous outcomes. Non-transferred embryos were partitioned into two groups: (1) Discarded – embryos that were excluded by the clinician because of defective morphology: poor morphology [8] or morophkinetics (i.e., slow or halted development). The discarded embryos were further partitioned into two subgroups: “blastocyst-discarded” (Video S2), embryos that reached blastulation, and “underdeveloped-discarded” (Videos S3-4), that did not reach blastulation. Based on these selection criteria we assume that discarded embryos fail to implant, and thus used these discarded embryos as a non-ambiguous label to assess the model’s performance when ranking KID-P over discarded embryos. (2) Frozen – visually proper embryos that were not selected for transfer because other sibling embryos from the same cohort were ranked higher. These embryos were frozen for a future transfer but were not considered for further analysis because their implantation potential was unknown. These partitions defined the embryo clinical ranking of implantation potential (Fig. 2A).

### Analysis

#### Cropping and resizing embryo images

A simple, low-cost, Convolutional Neural Network (CNN) was trained to segment and localize the embryo. The time-lapse incubation images are of size 500×500 pixels at seven focal planes. The segmentation network accepts a downscaled version of size of 128×128 pixels to enhance efficiency and speed. To deal with varying image focus we randomly selected different focal planes for training. The network outputs a pixel-wise binary classification segmentation mask of size 128×128. A U-NET architecture [28] was used, with 4 convolutional layers at each of the downlink/uplinks. Each layer was followed by a maxpool/upsample layer, ReLU activation function and batch normalization, where the number of features started from 8 and was doubled after each pooling. A diagram of the architecture and examples of the segmentation mask are shown in Fig. S1. Finally, the bounded embryo was cropped from the full sized (500×500 pixels) image and resized to 256×256 pixels image, denoted 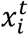.

#### Training classifier to discriminate KID-P embryos from different subsets of lower quality embryos

The data included the KID-P, KID-N, and discarded embryos that were split into Train/Validation/Test datasets (Tables S1). Each cohort of sibling embryos was allocated to one of the Train/Validation/Test datasets. Transferred embryos in the training set had incubation periods ranging from 2 to 6 days, whereas transferred embryos in the test set were composed solely of embryos that reached incubation durations of 114 to 120 hours to enable comparable results between different time points on the exact same set. This design makes the KID-N test set even more challenging compared to the training negative set since some of the embryos in the training set were transferred earlier on day 3 but would eventually have been labelled inviable, discarded and not transferred if they were allowed to incubate until day 5.

The learning task was to classify an input *x*^*t*^, where *x* is a feature representation of an embryo and *t* is time, to its ground-truth implantation binary outcome (detailed below). Features representations of an embryo included morphology (raw image), morphokinetic (timing of a predetermined set of embryo developmental stages) and each of these with the oocyte age as an additional feature. Details on each of these feature sets can be found below. The loss function used to train all three models was the binary cross entropy loss, a special case of equation (2), with K=2. The output of the classifier was a scalar 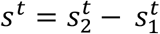 that represent the implantation probability *p*_*t*_:

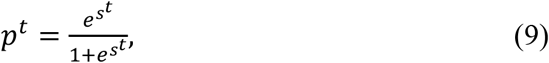

thus, all the models were of the form *F*: *x*^*t*^ → *s*^*t*^, where the feature representation *x*^*t*^ changed between models and *s*^*t*^ is the scalar prediction.

We trained twelve different classifiers. Six classifier where trained with morphology-based features and another six classifiers with morphokinetic-based features. Each classifier was trained to discriminate KID-P embryos from different sets of embryos (Table S2): (1) KID-P vs. KID-N; (2) KID-P vs. underdeveloped; (3) KID-P versus blastocyst discarded; (4) KID-P vs. all discarded; (5) KID-P vs. KID-N and all discarded; (6) KID-P vs. KID-P vs. KID-N and all discarded learned with oocyte age as an additional feature.

The morphology classifier, denoted Mh, received as input a single raw image. The morphokinetic classifier, denoted Mk, received a 15-dimensional vector holding the timing of specific morphokinetic events that were automatically extracted from the time-lapse stream. These two classifiers are described next.

#### Classification using morphological features

The morphology-based model *M*_*h*_ accepts single images as input and therefore exploits the embryo morphological information to predict implantation. The model was implemented using a CNN with a modified ResNet50 architecture [32]. To compensate for the drastic morphological changes throughout the embryo developmental process we used the ResNet50 base layers with multiple prediction heads 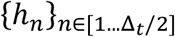, each corresponding to a temporal window of two hour. Each head consisted of a 128 channel dense layer, followed by a scalar output layer *s*^*t*^. Fig. S2A depicts the network’s architecture. The morphology-based classier was described in a separate manuscript that is currently under review and is attached to the submission.

#### Automating the timing identification of morphokinetic events with machine-learning and dynamic programming

The problem of estimating the timing of early morphokinetic events was studied recently up to four [29, 30], and up to eight [31] cells in the developing embryo, where all the later stages were considered as one state. We extended this scope to quantify both early and late morphokinetic events. Specifically, we automatically extracted timing of 15 morphokinetic events in the embryo development, i.e., the timing of specific morphological changes during embryo development identified from the time-lapse images (Fig. S3-4, Video S1). These morphokinetic features were then used to train classifiers (Fig. S2B).

More formally, the *morphokinetic state S* was defined by the following K = 15 morphokinetic events: Zygote, Pronuclei (PN) appearance and fading, 1Cell, 2Cell, |3Cell, 4Cells, 5Cells, 6Cells, 7Cells, 8Cells, 9^+^Cells, Morula, Start Blastulation, Blastocyst and Expanded Blastocyst.

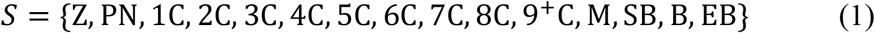

We combined machine-learning and dynamic programming to automated the timing identification of each specific morphokinetic event. The learning task was defined as classifying an image acquired at time *t, x*^*t*^, to its corresponding morphokinetic event *y*^*t*^ ∈ [1… *K*], where the ground-truth annotations were determined manually by an expert for model training and evaluation. The training data were a set of the form 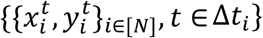, where N was the number of embryos, and Δ*t*_*i*_ was the time interval of the *i-th* embryo time-lapse incubation stream.

The morphokinetic model *M*_*k*_ is a multi-class classifier, trained using a CNN with a ResNet50 architecture [32] that accepts a single image 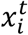 and predicts its morphokinetic event. 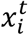 represents a cropped and resized image of the centered and resized embryo inside its well. The loss function was Categorical Cross Entropy loss [33] defined as

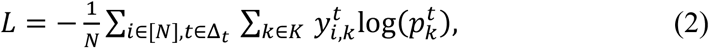

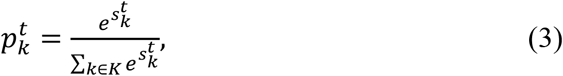

where 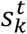 denotes the *k-th* output of *M*_*k*_ at time *t*. 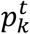 is derived from 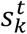 and represents the likelihood 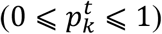 of each image *x*^*t*^ to be in *k* − *th* state. 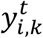 is the one-hot encoded ground-truth. Namely,

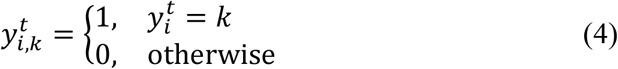

We denote by T, the set of K-1 morphokinetic events, defined by the starting time of all states in equation (1), except the first zygote, which is trivially defined by the time of the first frame 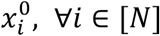. Namely,

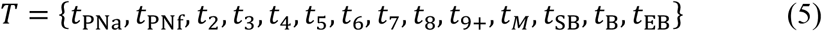

where *t*_*X*_ denotes the starting time of the X-th ev. ent

The K size vector *p*^*t*^ can be thought as a “soft annotation” per each frame. To estimate the sequence of morphokinetic events given the per-frame soft annotations, we used dynamic programming, where the updating step at time *t* + 1 is given by

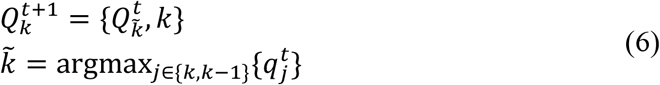

where 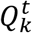 is a vector of size *t*, with the morphokinetic path at each time point *t* ∈ [1, *t*], that ends at the *k-th* state at time *t*, and 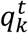 denotes a scalar score associated with the path 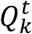. Where 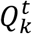 denotes the optimal path of state *k* at time *t*, in the sense that 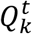 has the highest score 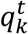 out of all possible paths that end at the *k-th* state at time *t*.

Further, the updated score 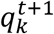 is given by

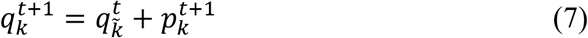

Simply stated, at each time *t* + 1, and for each state k, the optimal paths at time t of the states {*k* − 1, *k*} are compared. The best path of the *k-th* state at time *t* + 1 is the path with the highest score at time t, concatenated with the state *k* at time *t* + 1. The new path score is the sum of the score of the best path from the {*k* − 1, *k*} states and the score of the soft annotations score of the *k-th* state at time *t* + 1. Finally, the estimated ME vector ***m*** *=* [*m*_1_, *m*_2_, …. *m*_*k*−1_]^*T*^ is obtained by travelling over the transitions between states in 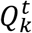. Namely:

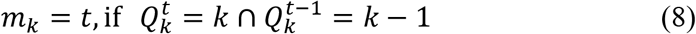

Fig. S3 demonstrates the soft annotations and dynamic programming outcomes for embryos with different developmental paths.

The accuracy of the annotation classification was measured by two measurements: (1) the average per frame accuracy, calculated as the average the 0-1 loss over frames, and (2) the confusion matrix of the morphokinetic events. Fig. S4 presents the confusion matrices of morphokinetic events for the morphokinetic classifier before and after applying the dynamic programming stage, where rows denote the ground truth annotations, and columns the multi-class prediction. The confusion rates were prominent only between adjacent events. The first and final events presented relatively low confusion rates, whereas the intermediate morphokinetic events were subject to greater confusion rates with adjacent events. For example, distinguishing between 5C and 6C is more difficult than between 1C and 3C or between the morula and start of blastulation. These results align with annotation errors by expert embryologists. One exception is a blastocyst embryo, which the model confused with expanded blastocyst. However, this can be explained by the relatively vague definition of expansion, which in turn led to an inter (and even intra) variability in annotating the exact timing of when expansion began. The confusion matrix for the dynamic programming decision executed over the multi-class prediction exhibited a higher true positive rate for all events and improved average accuracy to 85.5% compared to 83.6% for the multi-class prediction. This is attributed to the consideration of the full developmental trajectory as a global optimization problem.

#### Classification using morphokinetic features

The morphokinetic-based model *M*_*k*_ accepts as input the estimated ME vector ***m***, as calculated at time *t*. To allow the network to recognize embryos that stopped developing the time *t* was embedded to input vector. Formally, the input for model ***M***_***k***_ was the K-dimensional vector ***m***^*t*^, defined as

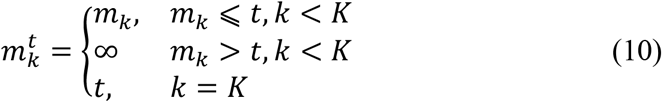

The classifier was implemented with a neural network that consisted of 6 consecutive dense layers. The first 5 layers with 256 channels, and the last hidden layer consisted of 128 channels. Each layer was followed by ReLU activation and batch normalization. The final output layer was the scalar prediction. Fig. S2B depicts the network’s architecture.

#### Classification using oocyte age

The oocyte age was determined by the age of the patient when the oocytes were retrieved from the ovary. Our dataset did not involve donors and did not include frozen embryos that were thawed and transferred. Thus, in this study, the oocyte age is equivalent to the patient/maternal age and is common to all embryos in a cohort. Oocyte age was previously reported to be highly correlated with implantation rates [18]. We included as an additional input to the morphology- and morphokinetic-based models, resulting in two new models denoted by ***M***_***ho***_, ***M***_***ko***_ respectively. The architecture of these models was identical to their original models with the addition of an embedded layer that accepted oocyte age, digitized by one-hot encoding to one of five possible age groups, and outputted 128 features that corresponded to the hot age group. This output was added to the last hidden layer of the original model, followed by a dense layer with 128 channels and a scalar output layer. Fig S2C depicts the network’s architecture.

#### Evaluating classifier performance in predicting embryo implantation and ranking

We evaluated the trained models’ performance with Area Under the Curve (AUC) measurement on four different binary classification tasks of discriminating KID-P embryos from different sets of embryos (Table S3): (1) KID-P vs. KID-N; (2) KID-P vs. all discarded embryos; (3) KID-P vs. blastocyst discarded; (4) KID-P vs. KID-N and all discarded embryos. This latter setting is most resembling the true embryo ranking in the clinic.

To directly assess the task of embryo ranking we evaluated the average number of attempts for embryo transfer from an IVF cycle until a successful implantation was reached, where the transfer order was determined by the model’s ranking based on its classification score. This measurement is called *time to live birth* (TTLB) [19]. This analysis included embryo cohorts that contained a successful implantation (KID-P) and multiple discarded embryos, where transferring a discarded embryo is assumed to result in a failed implantation. Thus, a basic requirement from a ranking model is to prioritize KID-P over discarded embryos. To avoid the bias of cohorts with different numbers of embryos we compared the average TTLB of sub-cohorts consisting of the same number of embryos sampled from the full cohorts. For a given number of embryos (N = 2-8) we created sub-cohorts by selecting the KID-P embryo and N-1 discarded embryos. We considered two scenarios: (1) calculating TTLB for sets of one KID-P and discarded embryos (blastocyst and underdeveloped); (2) calculating TTLB for sets of one KID-P and multiple blastocyst-discarded embryos (more challenging).

#### Software and data availability

We are currently organizing our source code and will make it publically available as soon as possible (before journal publications). The clinical data are owned by Hadassah Hebrew University Medical Center, the Soroka University Medical Center, and the NYU Langone Prelude Fertility Center. Patients’ data were anonymously used under ethical agreements with each clinic separately, without explicit patient consent for their data to be made public. Thus, restrictions apply to the availability of these data. Requests for the anonymized data should be made to Dr. Assaf Ben-Meir, Dr. Iris Har-Vardi, or Dr. James Grifo. Requests will be reviewed by a data access committee, taking into account the research proposal and intended use of the data. Requestors are required to sign a data-sharing agreement to ensure patients’ confidentiality is maintained prior to the release of any data. The methods presented are not specific to the datasets used in this study and users can train and test the deep learning model on any relevant imaging data.

## Funding and Acknowledgments

This research was supported by the Israel Council for Higher Education (CHE) via the Data Science Research Center, Ben-Gurion University of the Negev, Israel, by the Israel Science Foundation (grant No. 2516/21) and by the Welcome Leap Delta Tissue program (to AZ). We thank Maya Adani for designing and generating the figures. We thank Nadav Rappoport, Eran Eshed and Oded Rotem for critically reading the manuscript.

## Author Contribution

IE conceived the study, developed analytic tools, and analyzed the data. IE and AZ interpreted the data and drafted the manuscript. AZ mentored IE. All authors wrote and edited the manuscript and approved its content.

## Competing Financial Interests

IE, ABM and IHV are employees at Fairtility LTD. AZ is collaborating with AIVF LTD on projects not related to this study.

## Supplementary figures

**Figure S1:**
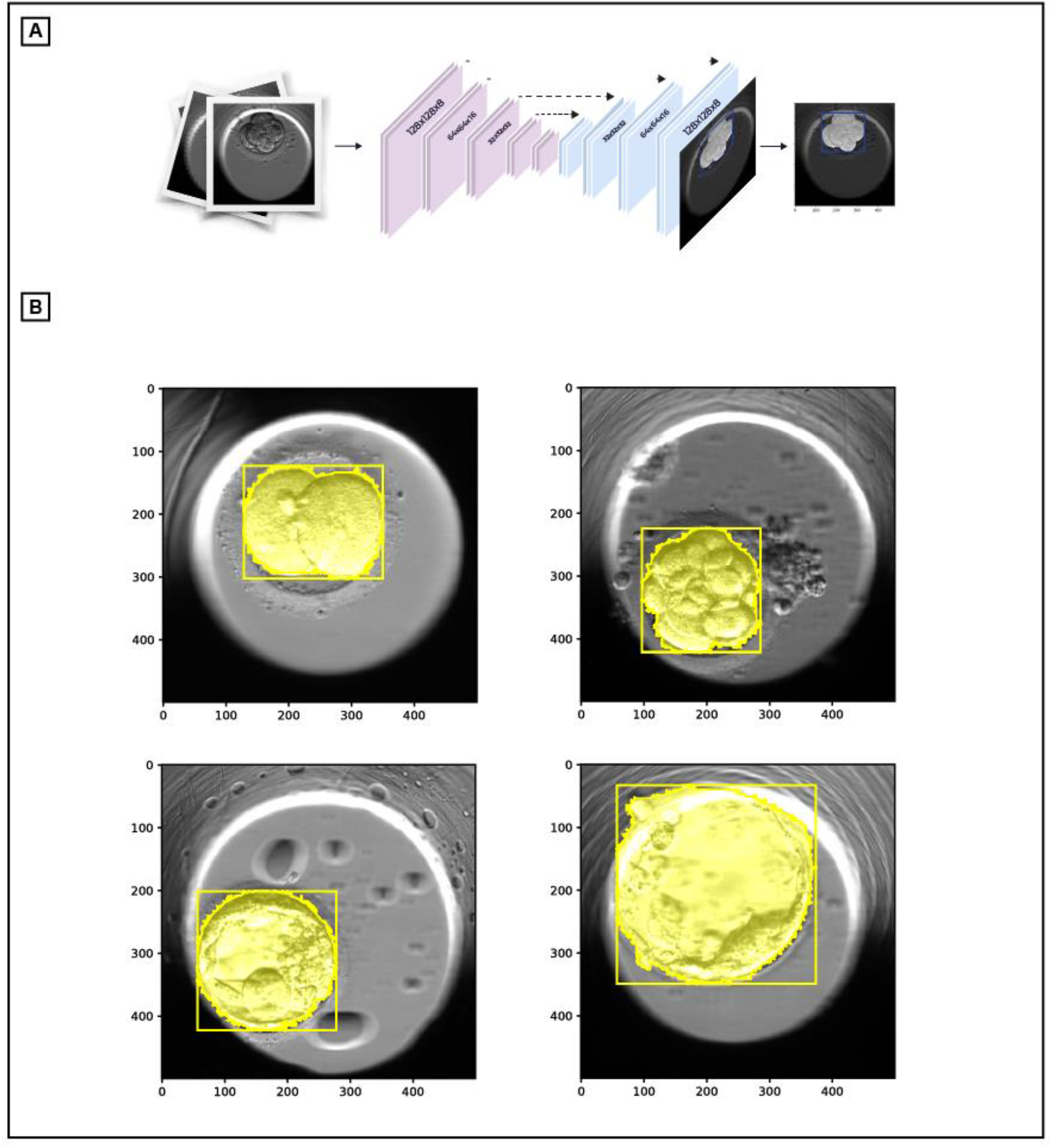
Embryo segmentation network. (**A**) U-NET architecture for embryo localization and segmentation. (**B**) Output masks for different developmental stage embryos. From upper left to lower right: 2 cells, 9+ cells, blastocyst, expanded blastocyst.

**Figure S2:**
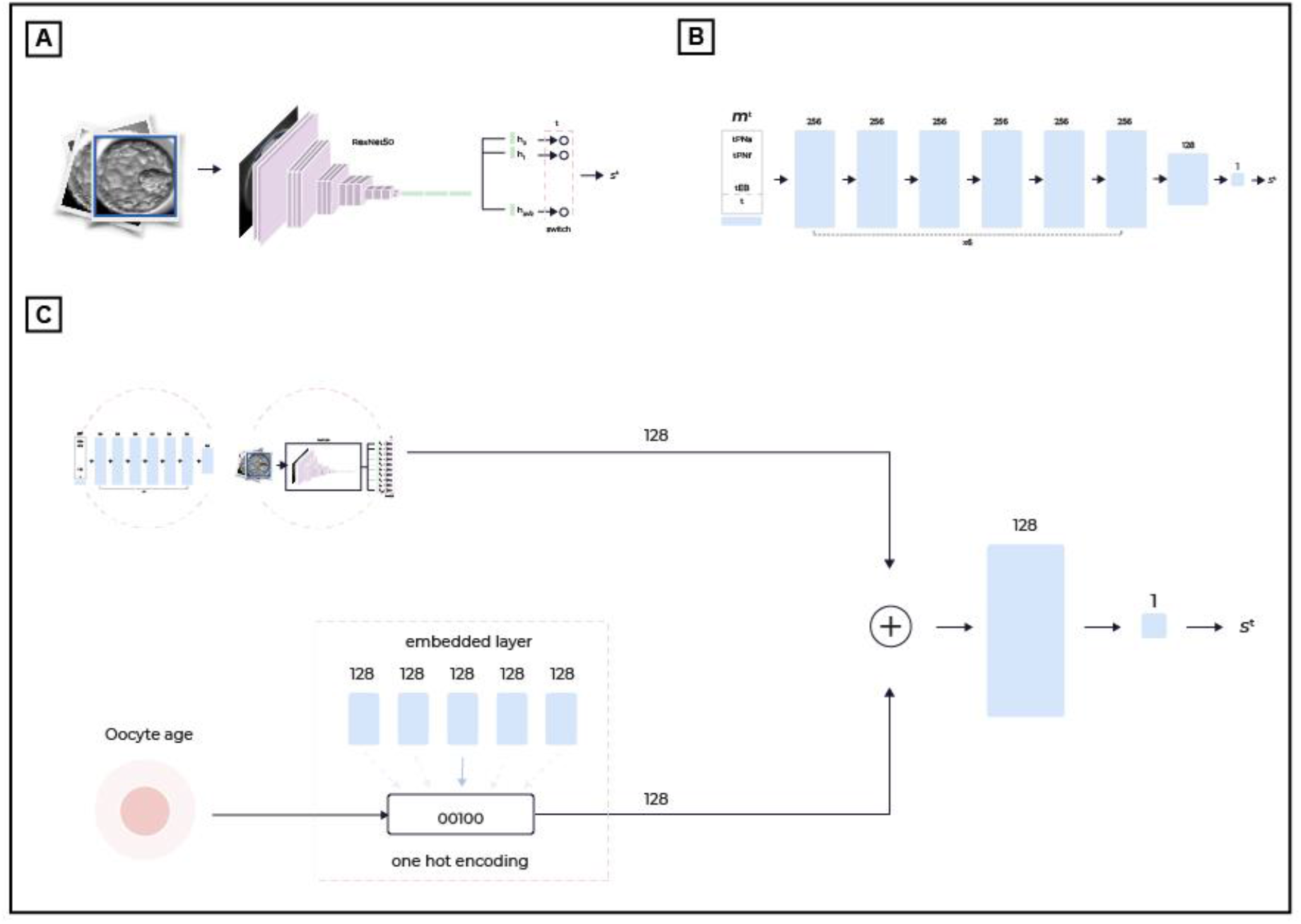
Implantation model architectures. (**A**) Morphological model *M*_*h*_, 128×128 input image, fed into Resnet50 CNN with multiple prediction heads. Each head corresponds to a temporal window of 2 hours. (**B**) Morphokinetic model *M*_*k*_, accepts vector ***m*** with the estimated starts of 15 morphokinetic events, and the time from fertilization. A simple neural network with eight dense layers was used. (**C**) The oocyte age was added as input to each model, resulting in models *M*_*ho*_, *M*_*ko*_ respectively. The oocyte age was passed through an embedded layer with 128 output channels. The embedding layer digitized the oocyte age to one of 5 age bins, and outputted the corresponding 128 channels. The embedded output was then added to the output of each of the basis models. Two dense layers conclude the architecture.

**Figure S3:**
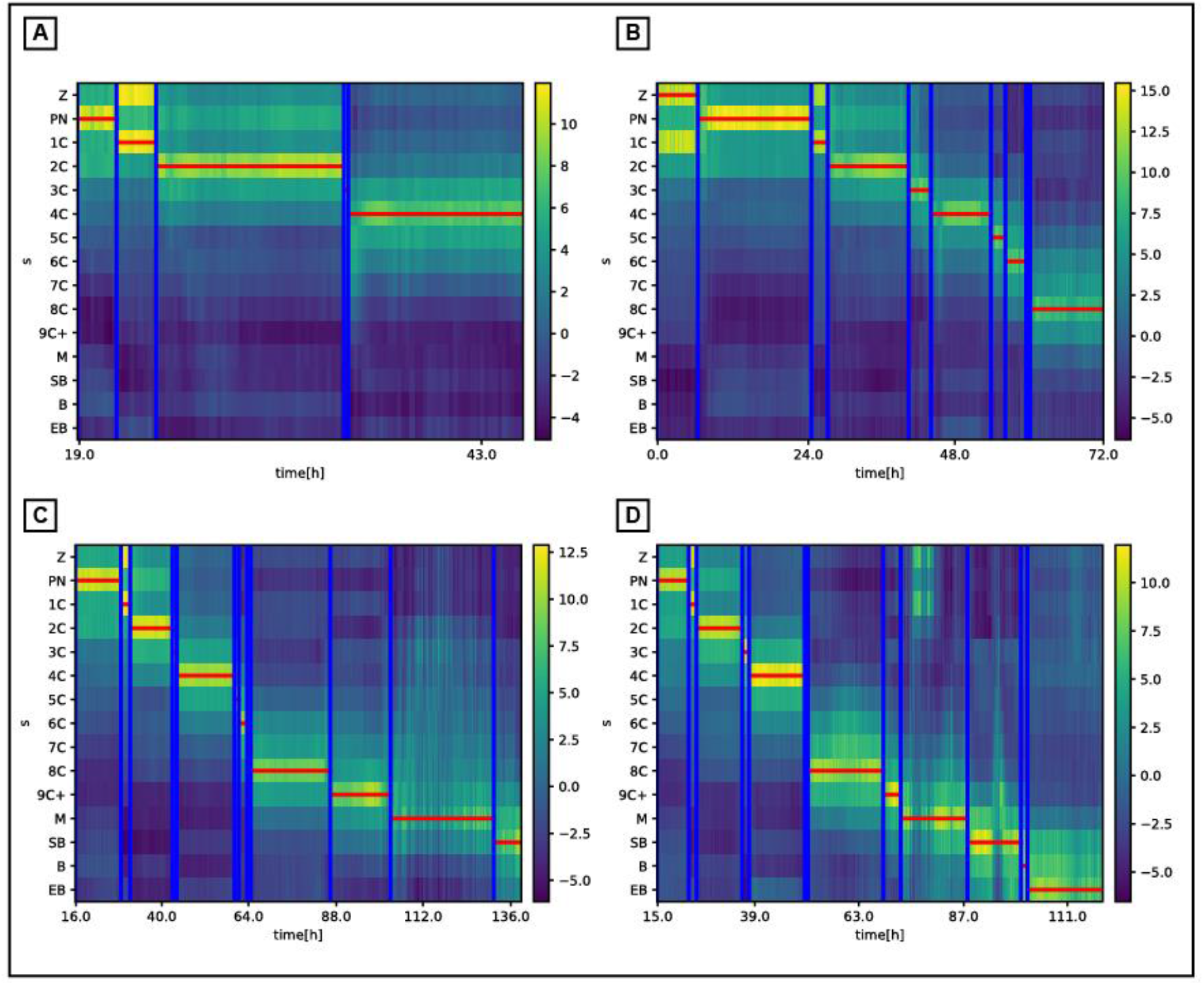
Predicted morphokinetic events vs. the ground truth (manual) annotations of different ending stage embryos: (**A**) 4Cells, (**B**) 8Cells, (**C**) Start Blastulation, (**D**) Expanded Blastocyst. The map represents the network multi-class prediction *s*^*t*^ of each frame vs. time. The horizontal lines are the dynamic programming output path 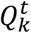, executed over the multi-class prediction map, and the vertical lines are the ground truth (manual) annotations. See Methods for full details.

**Figure S4:**
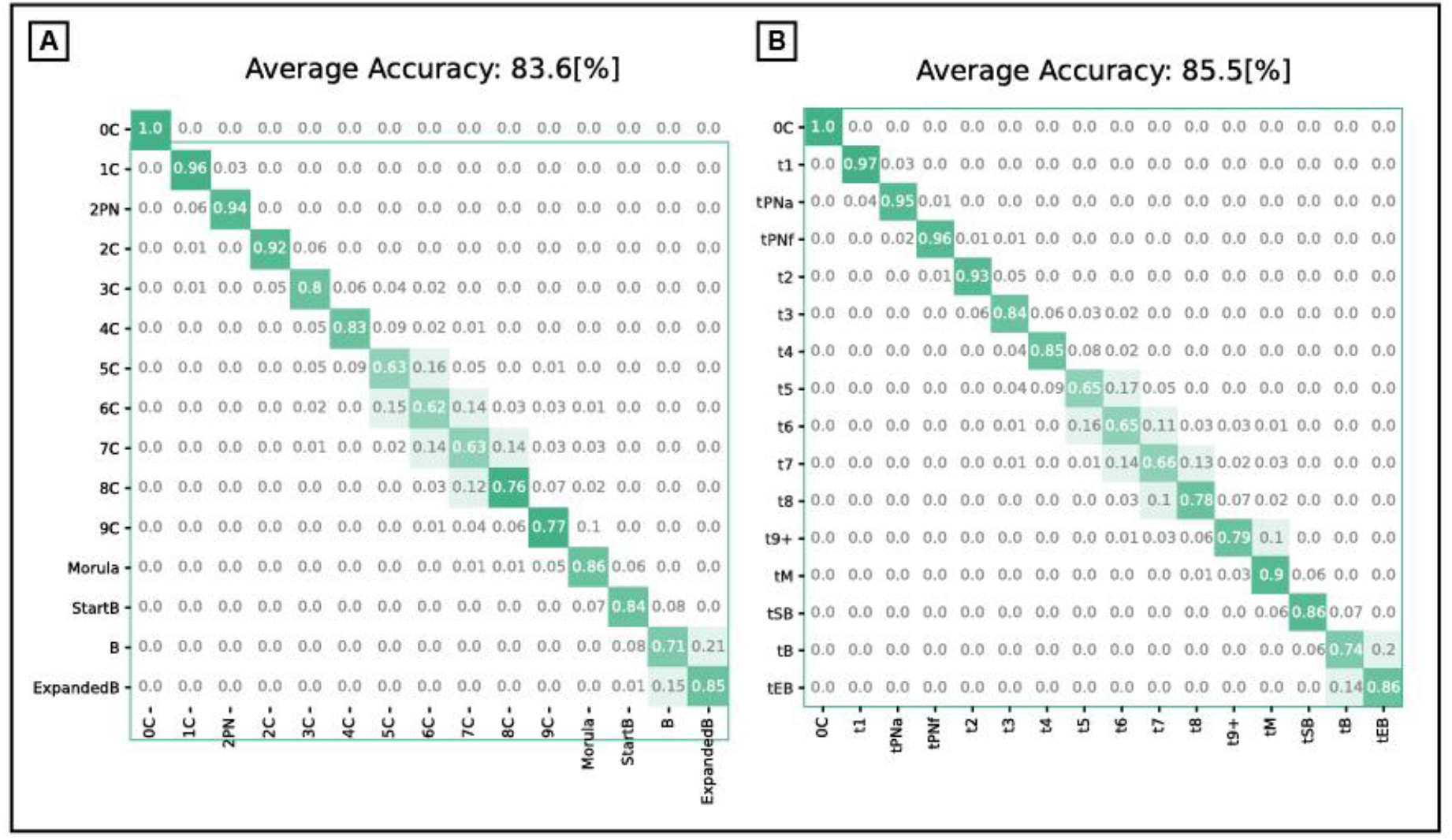
Confusion matrix of morphokinetic state classification. Rows denote the ground truth annotations, and columns the multi-class prediction. Shown are multi-class model predictions before (**A**), and after (**B**) dynamic programming.

**Figure S5:**
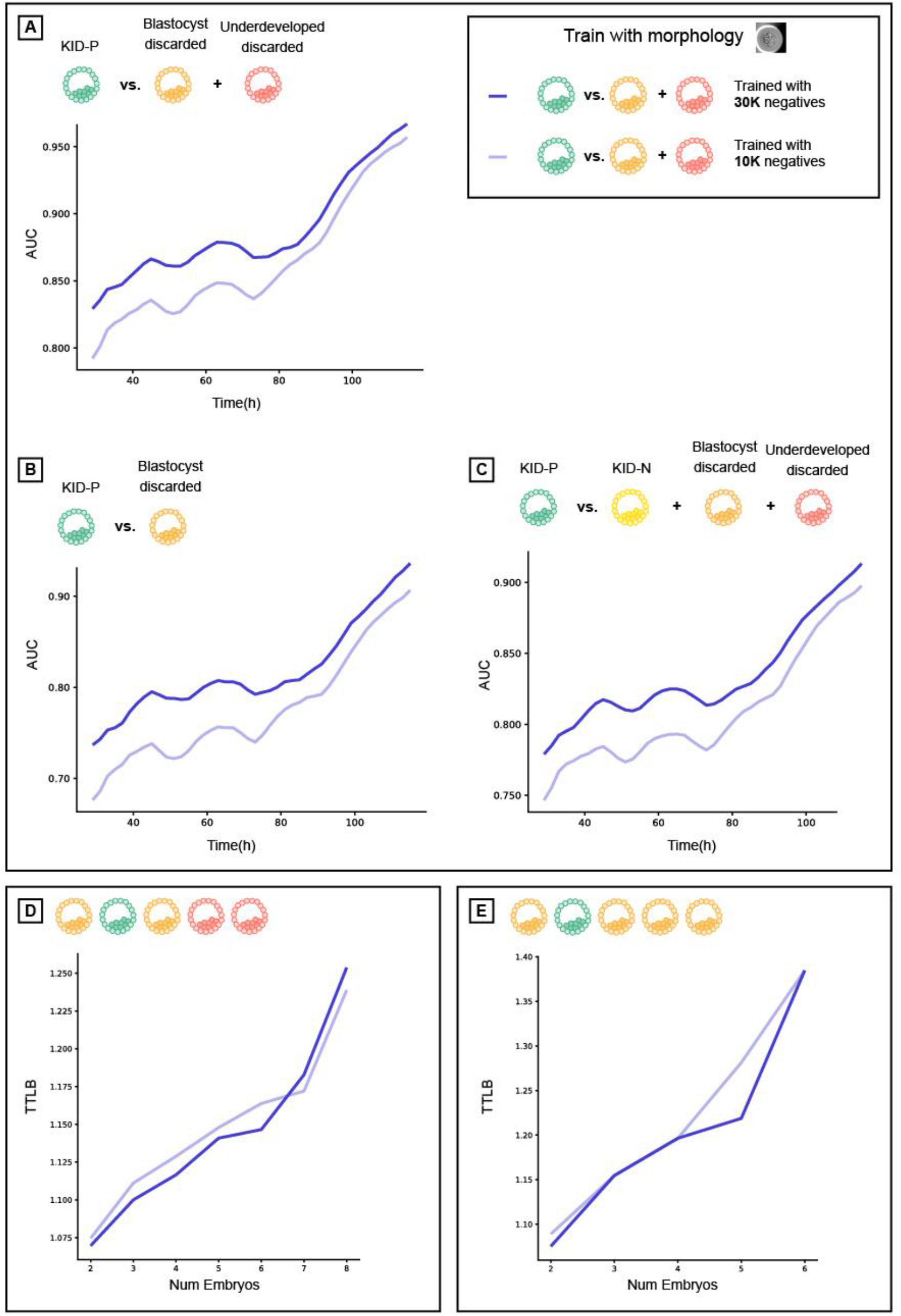
Additional training data enhances morphology-based models. Prediction accuracy: AUC vs. time since fertilization (hours) (**A-C**), average time to live birth (TTLB) (**D-E**) for morphology-based models, trained with different amounts of discarded embryos (negative labels): 10K and 30K, tested over different test sets. Training with more discarded embryos lead to better AUCs for all times and over all test sets, but did not change the TTLB. (**A**) KID-P vs. discarded. (**B**) KID-P vs. blastocyst discarded. (**C**) KID-P vs. KID-N and discarded. (**D**) Ranking KID-P and all discarded. (**E**) Ranking KID-P and blastocyst discarded.

**Figure S6:**
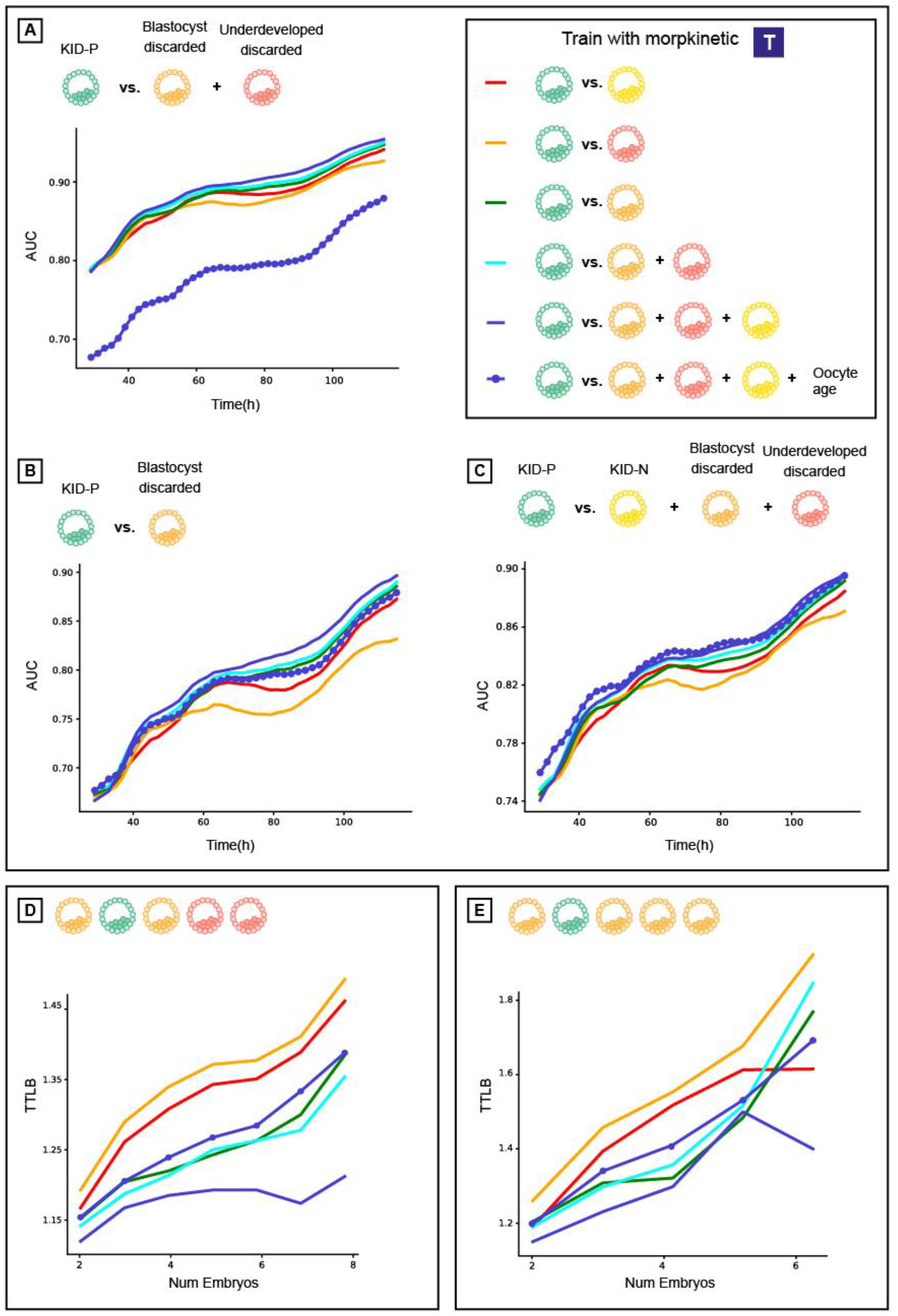
KID-N embryos are not the ideal negative label for solving the task of embryo ranking. Prediction accuracy: AUC vs. time since fertilization (hours) (**A-C**), average time to live birth (TTLB) (**D-E**) for morphokinetic-based models, trained over different negatives: KID-N, discarded, blastocyst discarded, underdeveloped discarded, and KID-N+discarded, tested over different test sets. (**A**) KID-P vs. all discarded. (**B**) KID-P. vs. blastocyst discarded. (**C**) KID-P vs. KID-N and all discarded. (**D**) Ranking KID-P and discarded. (**E**) Ranking KID-P and blastocyst discarded.

**Figure S7:**
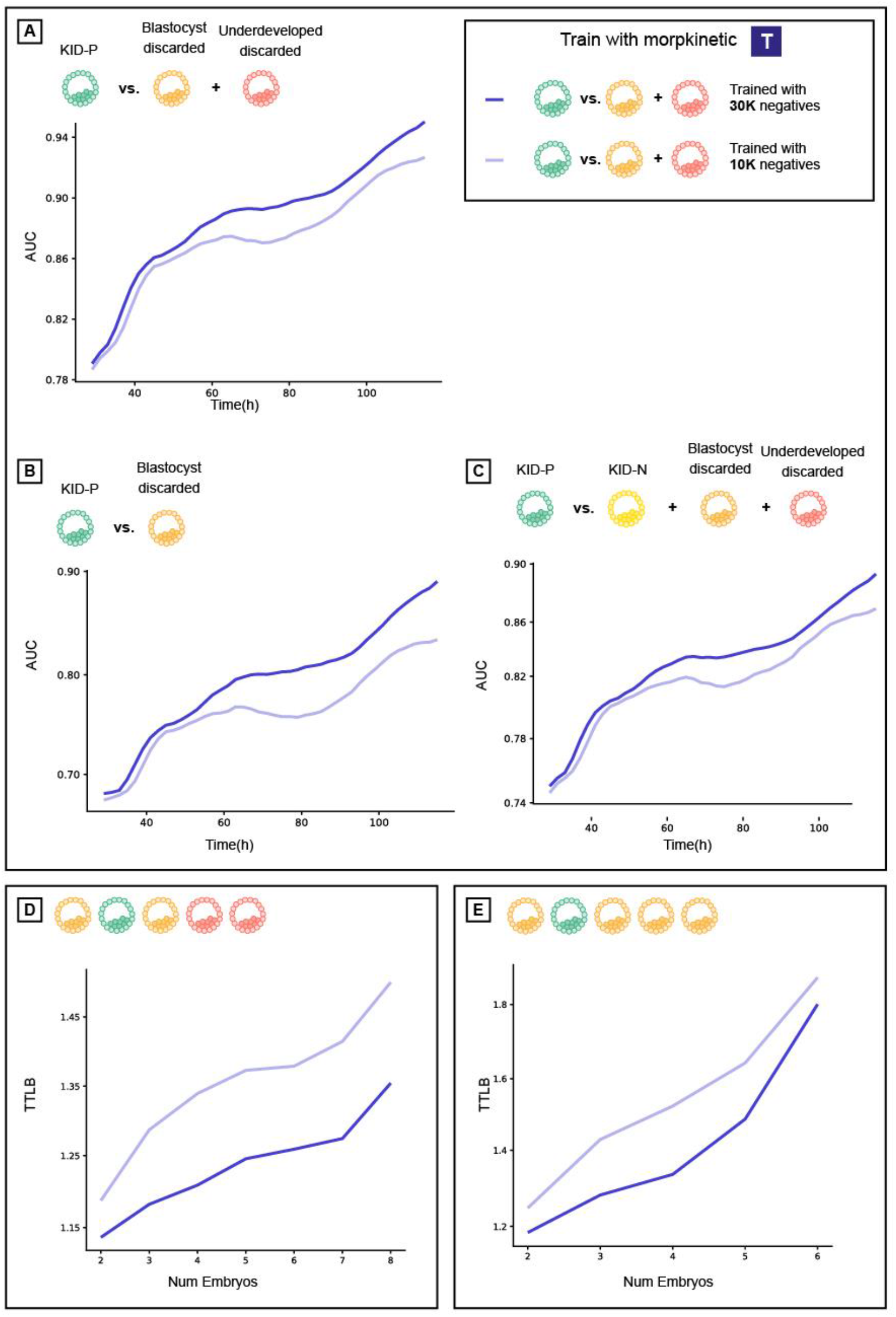
Additional training data enhances morphokinetic-based models. Prediction accuracy: AUC vs. time since fertilization (hours) (**A-C**), average time to live birth (TTLB) (**D-E**) for morphokinetic-based models, trained with different discarded amounts: 10K and 30K, tested over different test sets. Training with more negatives lead to better AUCs and yields to shorter TTLB over all test sets and times. (**A**) KID-P vs. discarded. (**B**) KID-P. vs. blastocyst discarded. (**C**) KID-P vs. KID-N and discarded. (**D**) Ranking KID-P and discarded. (**E**) Ranking KID-P and blastocyst discarded.

**Figure S8:**
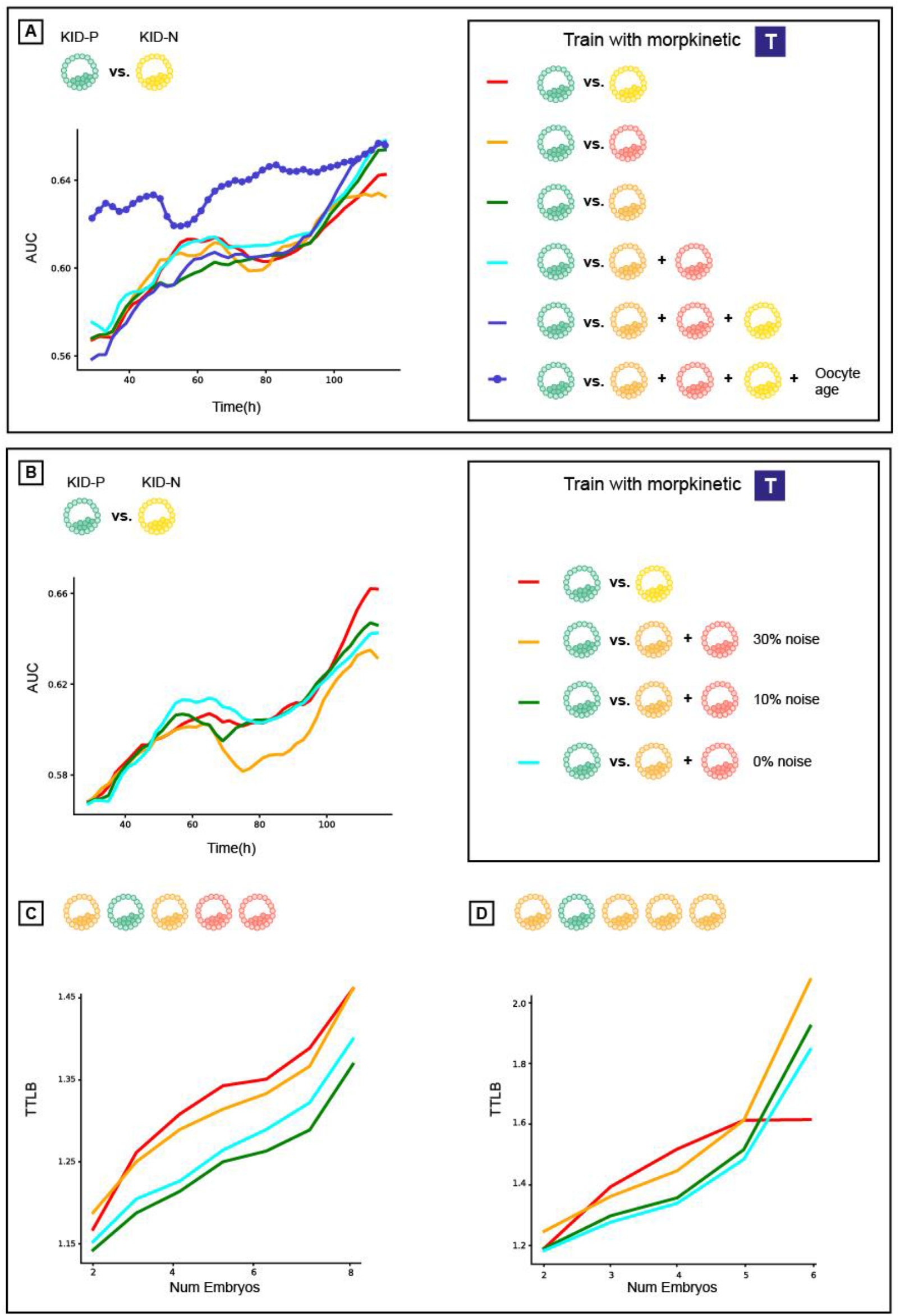
An analysis of optimizing implantation prediction accuracy by trading off label noise level and training data quality based on morphokinetic features. (**A**) Comparison of models trained on different negatives: KID-N, discarded, blastocyst discarded, underdeveloped discarded, and KID-N+discarded (with and without oocyte age) over KID-P vs. KID-N test set. (**B**) AUCs for models trained with known noise levels (of positive labels) over KID-P vs. KID-N test set (**C**) TTLB for models trained with known noise levels (of positive labels) over KID-P vs. discarded test set (**D**) TTLB for models trained with known noise levels (of positives labels) over KID-P vs. blastocyst discarded test set.

## Supplementary tables legends

**Table S1**. Data table. Divided into Train/Validation and Test sets, where all embryos from a given cycle where either used for train, validation or test.

**Table S2**. Train splits for the different trained models. The same amount of negative labels from each embryo discarded category was maintained across all sets of training in order to compare training with different negatives fairly. KID-N consists of the smallest amount of 6500 embryos, so 6500 embryos were randomly selected from all other negative sets

(Underdeveloped discarded + Blastocyst discarded, Blastocyst discarded, KID-N + Underdeveloped discarded + Blastocyst discarded and Underdeveloped discarded), first considering all valid negatives, and then selecting 6500 embryos at random from the valid set.

**Table S3**. Test splits for the different negatives sets.

## Supplementary videos legends

**Video S1**. Temporal evolution of a KID-P embryo. Bounding box marks the automatically detected embryo contour. Time (in hours) is displayed at the bottom-left. The predicted morphokinetic state and morphological-based implantation prediction score (KID-Score, from time = 30 hours) are displayed at the top-left. The KID-P score at the final frame is 1.

**Video S2**. Temporal evolution of a blastocyst discarded embryo. Bounding box marks the automatically detected embryo contour. Time (in hours) is displayed at the bottom-left. The predicted morphokinetic state and morphological-based implantation prediction score (KID-Score, from time = 30 hours) are displayed at the top-left. The embryo was discarded after 126 hours, reaching the blastocyst (tB) stage, with a KID-P score of −0.95.

**Video S3**. Temporal evolution of an underdeveloped discarded embryo. Bounding box marks the automatically detected embryo contour. Time (in hours) is displayed at the bottom-left. The predicted morphokinetic state and morphological-based implantation prediction score (KID-Score, from time = 30 hours) are displayed at the top-left. The embryo was discarded after 127 hours, reaching the start of blastulation (tSB) stage with a KID-P score of −1.31.

**Video S4**. Temporal evolution of a very low-quality underdeveloped discarded embryo. Bounding box marks the automatically detected embryo contour. Time (in hours) is displayed at the bottom-left. The predicted morphokinetic state and morphological-based implantation prediction score (KID-Score, from time = 30 hours) are displayed at the top-left. The embryo stopped developing after reaching the Pronuclei fading stage (tPNf), with a KID-P score of −9.

## Notes

### Author Declarations

Human embryo image/video data collected from patients were used in this study with institutional review board approval from the Investigation Review Board of Hadassah Hebrew University Medical Center (IRB# HMO-006-20)

